# Covid-19 Incidence Rate Evolution Modeling using Dual Wave Gaussian-Lorentzian Composite Functions

**DOI:** 10.1101/2020.06.07.20124966

**Authors:** Radhakrishnan Poomari

## Abstract

Modeling the evolution of Covid-19 incidence rate is critical to deciding and assessing non-medical intervention strategies that can lead to successful containment of the pandemic. This research presents a mathematical model to empirically assess measures related to various pandemic containment strategies, their similarities and a probabilistic estimate on the evolution of Covid-19 incidence rates. The model is built on the principle that, the exponential rise and decay of the number of confirmed Covid-19 infections can be construed as a set of concurrent non-linear waves. These waves can be characterized by a linear combination of Gaussian and Cauchy Lorentz functions collectively termed as Gaussian-Lorentzian Composite (GLC) function. The GLC function is used for non-linear approximation of officially confirmed Covid-19 incidence rates in each country. Results of fitting GLC based models to incidence rate trends of 20 different countries proves that the models can empirically explain the growth and decay trajectory Covid-19 infections in a given population.

---

The World Health Organization declared Covid-19 outbreak as a global pandemic on March 11, 2020 [1]. Since then, a number of Covid-19 epidemic spread models were developed using data officially reported by countries across the globe. It is well known that current data on Covid-19 confirmed infections does not reflect the true scale of spread of the pandemic. However, a wide variety of forecasting models have been developed based on SIR deterministic framework and its variants given their historic significance in epidemic modeling [2, 3, 4, 5, 6].

One of the most fundamental metric used by the various SIR based deterministic frameworks is the basic reproduction number, *R*_0_ which is used to quantify the threshold of an epidemic outbreak. The *R*_0_ number defined as expected number of secondary cases that can be produced by a single infected individual in a completely susceptible population, is estimated based on assumptions on transmissibility, contact dynamics and expected duration of infection. The calculation, usage and interpretation of *R*_0_ in a model may lead to flaws unless the model explicity defines the base assumptions, limitations and source of information [7]. Several studies have highlighted key challenges for epidemic modeling when using deterministic, stochastic and network based methods that requires estimation of *R*_0_ to be able to forecast the evolution of the epidemic spread [8, 9, 10, 11]. A wide spectrum of factors such as Genetic diversity, Population Immunity, Mobility Pattern, Social contact dynamics, Spatial structures, Pathogen mutation and evolution, Exposure time to pathogen, Viral load, Super spreaders, Seasonality and Endemicity introduce large scale uncertainties. It is near impossible to collect enough and qualitative data regarding each of the forementioned factors and develop a generic model, factoring in all uncertainties and make a forecast on epidemic spread.

In search for a simple generic epidemic model, there have been studies published on using empirical mathematical functions for characterizing the spread of an epidemic based on the trajectory of its curve. An inverted parabola function was used to approximate single-wave epidemic outbreaks based on cumulative number of incidence rates [12]. The inverted parabola model was tested on nine different epidemic outbreaks across various geographies that include Chikungunya, Ebola, Salmonella SaintPaul, Gastroenteritis, Pertussis to forecast future cumulative number incidences. Another approach leveraged a sub-epidemic wave model that considers the trajectory of an epidemic, its peaks and the variations in its oscillations to make a short term forecast [13]. The sub-epidemic wave model was based on a generalized logistic growth model (GLM) function based on the study [14] that was originally applied to outbreak data on SARS in Singapore, Plague in Madagascar and Ebola in the Democratic Republic of Congo. The sub-epidemic wave model was shown to perform well for short term forecasts of two to three weeks and when the incidence pattern was stable. However, when there was significant increase in incidence rate the model was unable to capture the pattern within certain components of sub-epidemic waves. Considering the current Covid-19 pandemic, the scale of testing across a given population and the reporting delays of incidence rates introduce substantial uncertainty. These complexities along with varying non-medical interventions and policies by different Government organizations at both the provincial and federal levels, makes it almost impossible to gauge the exact number of infections existing in a populations leading to a highly non-linear time varying pandemic wave.

Acknowledging the best attempts made by various studies in decoding the trajectory of the Covid-19 pandemic, there is still a need for a method that has less complexity, encode the wide variety of uncertainties that influcence incidence rates and can be generically applied to data across any geographical region. Most importantly, the method should be able to account for high variations of officially reported incidence rates and untested asymptomatic population considering a potentially endemic characteristic of Covid-19. This paper presents a mathematical function that can model the current Covid-19 pandemic’s growth and decay trajectories based on the reported incidence rates from each geographical region. The results of the model compares the evolutionary trend of incidence rates across 20 different countries with the discussion on the various non-medical interventions by the corresponding Governments and its citizens’ response and cooperation.

## Materials and Methods

### Data Sources

The data used for this research was obtained from the COVID-19 Data Repository by the Center for Systems Science and Engineering (CSSE) at Johns Hopkins University [15]. Specifically, the time series dataset for global confirmed cases was used for the mathematical modeling. The time series dataset is a daily updated data file with incidence rate numbers from each country from where official data was made available. For the purpose of analysis, the daily time series dataset was transformed to a weekly time series dataset. This provides the ability to visualize and gain insights into the weekly incidence rate growth and decay with respect to time. Also the reported incidence numbers do not reflect the actual reality of infection spread due to the fact the patients who got tested for Covid-19 had their results available only after a certain time period.

### Mathematical Modeling

The Covid-19 incidence rate growth and decay and be construed as non-linear wave traveling through time. In order to model the non-linearity we propose a non-linear approximation model using a linear combination of Gaussian and Cauchy Lorentz composite functions. The Gaussian and Lorentz composite functions have been earlier introduced and applied to surface chemical analysis and peak fitting of X-ray photoelectron spectroscopy (XPS) data [16]. Both Gaussian and Cauchy Lorentzian functions have specific properties which when used in combination as a composite function can be used to linearly approximate any non-linear wave. The following general mathematical expressions Eqs. (1a), (1b) and (1c) respectively define the Gaussian, Lorentzian and GLC function.

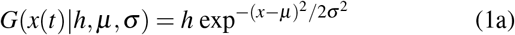

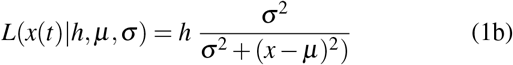

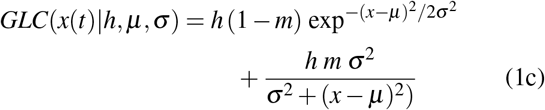

*x*(*t*) = weekly Covid-19 incidence rate

*h* = amplitude indicating the peak value of weekly incidence rate

*µ* = mean value indicating the time period when the incidence rate peaked

*σ* = standard devation characterizing the variance in the incidence rates across the time

In the above equations, the factor m is a constant that blends both the Gaussian and Lorentzian function into the GLC function. When *m* = 0, (1c) reduces to (1a) an when *m* = 1, (1c) reduces to (1b).

The mathematical functions defined by (1a), (1b) and (1c) are illustrated by Figure.1. The graph is plotted based on Covid-19 incidence data for Taiwan. The parameter values used to compute the functions for each of the three curves are *h*=144, *µ*=64 and *σ* =13.7. It can be noted that the Gaussian function represented by the green curve starts its growth from zero and ends at zero indicating that the origin and end of the pandemic are at specific time periods. The issue with modeling incidence rate evolution with a pure Gaussian function is that it does not take into account the uncertainty of when the actual spread of Covid-19 infection started in a population. Since data was available from the Jan 22, 2020, it is assumed that incidences from that particular day onward marked the starting of the outbreak. However, in reality it is a well known fact the timeline of spread of infections in a population is unknown and only when Governments begin testing a small fraction of the population, the officially reported numbers become the baseline for all modeling efforts. Likewise, the Covid-19 pandemic decay trajectory does not end towards zero incidences and it continues to linger among the population indicating a possibility of becoming endemic [17].

**Figure 1.**
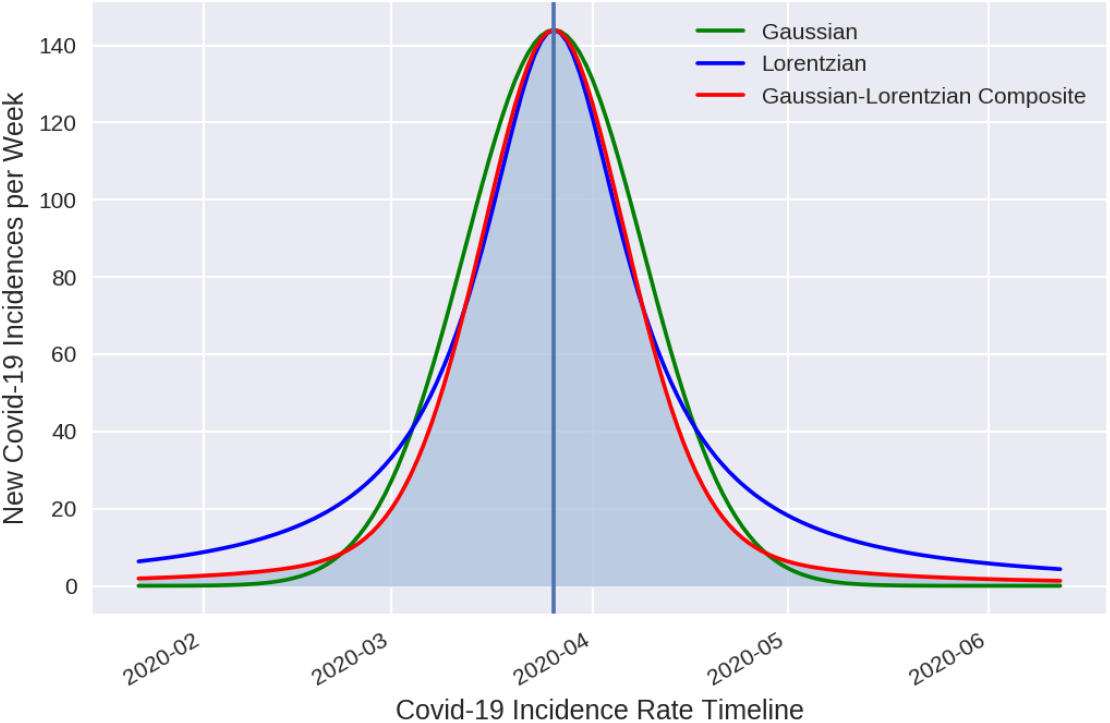
Gaussian, Lorentzian and GLC Curves

Despite its direct inapplicability towards modeling the evolution of Covid-19 incidence rate, the Gaussian function has a unique property defined by the Central Limit Theorem (CLT). The CLT states that when an observed random variable is the sum of many random processes, the resulting distribution of the random variable is a Gaussian distribution. Further, the Gaussian function has been used for non-linear approximation problems due to its simplicity and mathematical transformative properties [18, 19]. To account for the uncertainties during the start, growth, decay and the end of the Covid-19 incidence rates, the Lorentzian function is chosen. The distribution defined by a Lorentzian function is also referred to as a pathological distribution since its expected value and mean are undefined. Referring back to Figure.1, the blue curve represents the Lorentzian function. The red curve representing the GLC function has the mathematical properties of both the Gaussian and Lorentzian functions to model the Covid-19 incidence rate evolution accounting for high non-linearity and uncertainty.

Upon analyzing the Covid-19 incidence rates for 20 countries, the evolutionary trajectory of weekly incidence rates which is being considered as a non-linear wave can be linearly approximated using two GLC functions at different scales. More specifically, we consider a pair of GLC wave components in a linear combination to approximate the non-linear incidence rate trend. Using (1c), the dual wave GLC function can be formulated as follows.

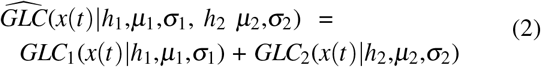

The GLC function and the dual wave GLC functions are illustrated in Figure. 2. Referring to Figure.2, the green and orange curves represent the GLC model for first and second wave components respectively for Taiwan. The first wave component represents the evolutionary phase of Covid-19 weekly incidence rate wherein a clear trajectory is evident indicating the start, growth, peaking and decay beginning from the time period Jan 22, 2020 from when data is currently available. Further, it can be noticed that around April 22, 2020, there seems to be sudden a surge in the number of weekly incidences. This phenomenon indicates a second wave component which has its own start, growth, peaking and decay trend. Mathematically, the probability density functions of the GLC curves denote the likelihood of measuring a particular value of the Covid-19 weekly incidence rate at a specific point in time. However, compared to the first wave component, the amplitude or peak of the second wave component is much smaller.

**Figure 2.**
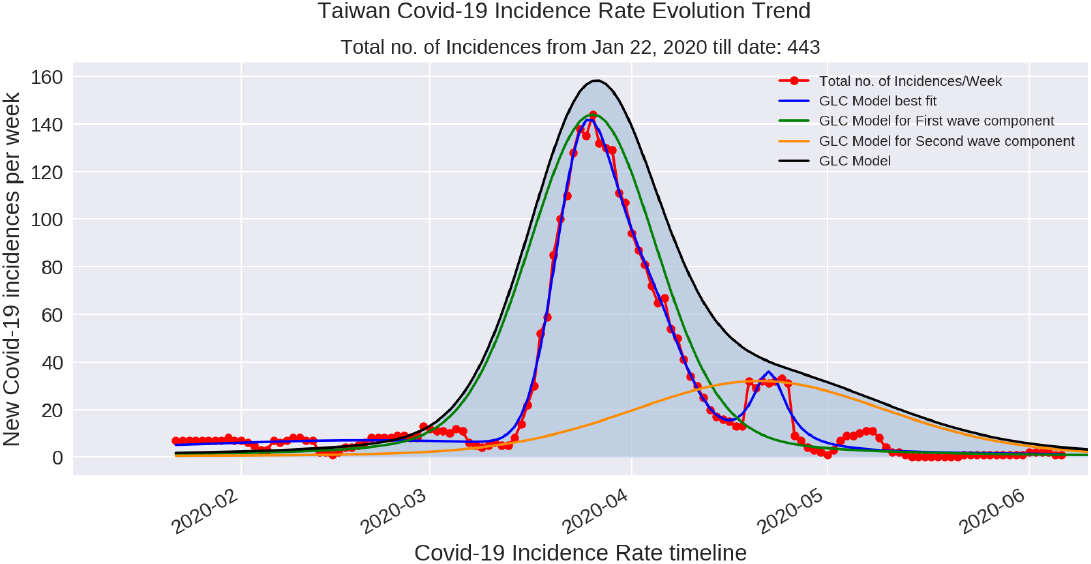
Incidence Rate Trend Model for Taiwan wherein the second wave component is smaller than the first wave component.

The second wave component is introduced in order to account for the dynamics of resurrgence of Covid-19 incidences or the outcome of non-medical Governement interventions in a population to contain the spread of the pandemic. Two specific hypothesis are considered that can explain the occurrence of the second wave component. First, it can be attributed to the low number of tests for Covid-19 infections done on a small fraction of a country’s population. Due to the low testing rates, the segment of the population who were asymptomatic earlier may start showing symptoms for Covid-19 later and by then they could have further spread the infection in the community potentially contributing to the weekly incidence numbers. The second hypothesis is that in inspite of non-medical interventions such as local or national lockdown strategies of the Governments, the compliance of citizens to various policies to contain the spread of Covid-19 cannot by fully ascertained and enforced across the entire population. In some countries, the size of the population, socio-economic structure, population density, cultural factors and mobility dynamics can contribute to non-compliance towards non-medical interventions. Considering both the hypothesis, it can be inferred that there is a likelihood of presence of a second wave component that can be smaller or larger compared to the first wave component.

Figure.3 illustrates the incident rate trend model for Hong Kong wherein the second wave component is larger than the first. The dual wave GLC function represented by the black curves in both Figure. 2 and Figure. 3 is a non-linear approximation of both first and second wave component. The best fit model of dual wave GLC function to weekly Covid-19 incidence rate is illustrated by the blue curve. The best model fit is obtained using a combination of Nelder-Mead method and Levenberg–Marquardt’s Damped Least-Squares method. The best fit model is presented in this paper only to highlight that mathematical models trying to deliberately perform a best fit on the existing data are highly biased. As stated in the beginning of the paper, the officially reported, Covid-19 incidence data does not reflect true spread of the pandemic and hence the best fit model of any method is fundamentally flawed. On the contrary, the non-linear approximation of both the first and second wave components by the dual wave GLC function is an optimal solution to modeling the evolution of Covid-19 incidence rates. Futher, the probability density of the dual wave GLC function also can account for the uncertainty and variance in incidence rate evolution over a defined time period.

**Figure 3.**
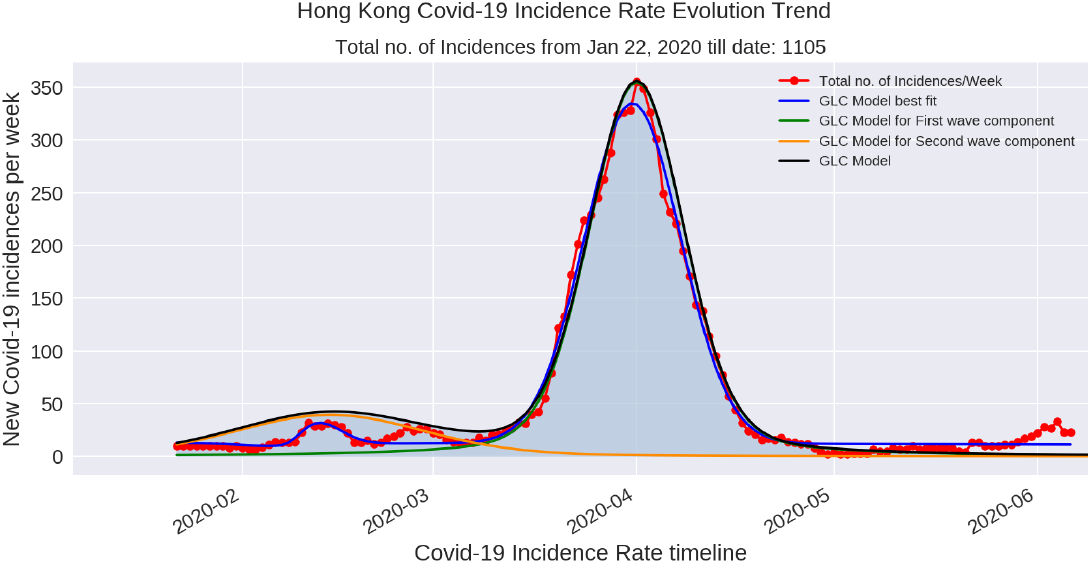
Incidence Rate Trend Model for Hong Kong wherein the second wave component is larger than the first wave component.

### Parameter Estimation

Estimating the parameters of the GLC function is critical in effectively approximating the non-linearity of Covid-19 incidence rates. Parameter estimation is done based on the following series of mathematical relationships of first and second wave components iteratively.

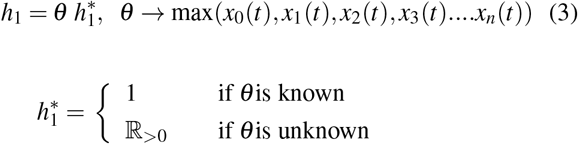

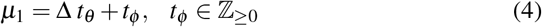

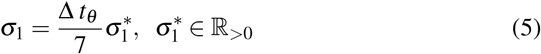

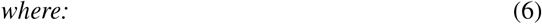

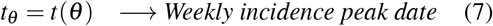

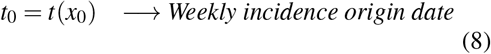

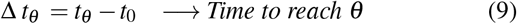

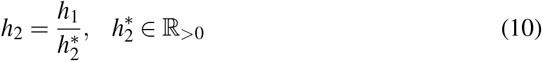

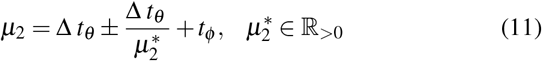

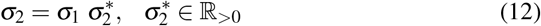

Availability of enough and quality data is highly critical to any mathematical analysis. Trying to model a highly nonlinear phenomenon like Covid-19 incidence rate evolution at the early stage wherein data is sparse can lead to biased outcomes that can negatively influence strategic decisions on epidemic management. Hence it is prudent to wait for the availability of the right amount of data to be able to model and understand fundamental patterns of infection spread in a population. One of the key requirements of the GLC method is the availability of incidence rate data on the complete evolutionary trend atleast for a set of geographic regions. In this research, the dual wave GLC function was initially applied to Taiwan, Hong Kong, South Korea, Australia, New Zealand since their incidence rate data had the complete trajectory of the curve encoded from start to flattening of the curve. Parameters were estimated for the first and second wave components of each of the forementioned countries by iterative greedy selection of the value of the constants 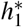, *t*_*ϕ*_ 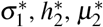 and 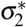 detailed in Table 1.

**Table 1.** Constant values for dual wave GLC parameter estimation. The values of the constants were chosen based on Global Covid-19 incidence data as on May 31, 2020.

|  | First Wave Component |  | Second Wave Component |  |  |  |
| --- | --- | --- | --- | --- | --- | --- |
| | $h_1^*$ | $\sigma_1^*$ | $\mu_2^*$ | $h_2^*$ | $\sigma_2^*$ | $t_\phi$ |
| Australia | 1 | 1 | 4 | 9 | 2.5 | 0 |
| New Zealand | 1 | 0.8 | 6 | 6 | 2 | 0 |
| Taiwan | 1 | 1.5 | 2.5 | 4.5 | 2 | 0 |
| Hong Kong | 1 | 1 | 1.5 | 9.0 | 2.0 | 0 |
| South Korea | 1 | 1 | 2 | 4.5 | 2.5 | 0 |
| Japan | 1 | 1.5 | 2.5 | 11 | 1 | 0 |
| Norway | 1 | 1.5 | 4.5 | 2.8 | 2.5 | 0 |
| Spain | 1 | 1.5 | 4 | 1.7 | 2 | 2 |
| Germany | 1 | 1.5 | 5 | 2.2 | 2 | 2 |
| France | 1 | 1.2 | 5 | 1.8 | 0.9 | 4 |
| Italy | 1 | 1.5 | 4 | 1.5 | 2.5 | 4 |
| Singapore | 1 | 1 | 5.5 | 1.3 | 2.5 | 0 |
| US | 1 | 2 | 5.5 | 1.1 | 4.0 | 0 |
| UK | 1 | 1.5 | 4.5 | 1.1 | 2 | 4 |
| Russia | 1 | 1.5 | 5.5 | 1.2 | 2.5 | 2 |
| Canada | 1 | 1.8 | 3.5 | 1.6 | 1.4 | 4 |
| UAE | 1 | 1.6 | 5.5 | 1.5 | 1.0 | 0 |
| India | 1.5 | 2.0 | 5.5 | 1.5 | 2.2 | 25 |
| Sweden | 1.5 | 2.5 | 2.0 | 2.0 | 0.8 | 28 |
| Brazil | 1.4 | 2 | 5.5 | 1.1 | 2.2 | 20 |

Referring to (3) and (10), 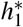 and 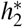 represents scaling factors that are used to control the amplitude of the first and second wave components respectively. 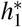 is defaulted to 1, if *θ* which indicates the maximum value of weekly incidences can be estimated based on available data. Hence for countries that have reached their peak incidence rate and have started showing a decaying trajectory, 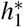 will be 1. Analyzing the estimated amplitudes of *h*_1_ and *h*_2_ for each of the 20 countries reveals that undetected spread of infection prevalent in a population and the low testing rates manifest as the second wave component after the first wave component reaches its peak. This explains the asymmetrical nature of the incidence rate curve wherein the decay trajectory has a longer oscillatory tail compared to the exponential growth trend at earlier phase of the curve. In a majority of the cases, the value of *h*_1_ is higher than *h*_2_. However, in some cases like Hong Kong, Japan, Finland, France and UAE, the amplitude of *h*_1_ is smaller than the amplitude of *h*_2_.

As on current date of this research, India, Brazil and Sweden have not yet reached their peak of weekly incidence rates. In such cases, the 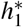 value need to be chosen accordingly to account for their growth trajectory and when they may reach their peaks. The choice of the 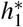 value is made by comparing the similarity of incidence rate evolutionary trajectories of other countries who have successfully flattened the curve along with other factors such as Covid-19 positivity rate and population to testing rate estimated based on public data source [20]. Further, 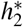 is selected according to the trajectory of the first wave component whether it is yet to reach the peak or has reached the peak and is on its decay trend. The scaling factor 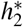 creates an inverse relationship between *h*_1_ and *h*_2_.

The constant *t*_*ϕ*_ in (4) is chosen to quantitatively account for the effects of non-medical interventions such as lockdown and recommendation policies by Governments across each country. Non-medical interventions range from national or local recommendations on Covid-19 health protocols to strict enforcement of local or national lockdown of economies [21]. Upon analyzing Table.1, it can be noted that countries that had strict local or national lockdown measures have a value of *t*_*ϕ*_ *>* 0. A non-zero value of *t*_*ϕ*_ results in shifting of the center *µ*_1_ of the first wave component with respect to the start of incidence reporting date. This means that any strict measures enforced to restrict mobility and economic activity of a population resulted in shifting of the amplitude or the peak *h*_1_ of the incidence rate evolution trajectory by a factor of *t*_*ϕ*_ days.

For example, the value of *t*_*ϕ*_ for Sweden, India and Brazil are much higher compared to any other country. Hence it can be inferred that the national and local lockdown measures in India and Brazil have postponed the date on which the incidence rate trend curve will reach its peak. India’s national lockdown started on Mar 24, 2020 and further enforced a series of extended lockdowns till the end of May 2020. Till May 2020, India has not reached the peak of its weekly incidence rate trend. Countries that have a smaller value of *t*_*ϕ*_ reached their peak of incidence rate evolution after enforcing strict lockdown measures but at the cost of very high incidence and mortality rates. On the contrary, Sweden is one of the countries which never had strict lockdown measures and has not yet reached the peak of incident rate curve at the time of this study. Therefore it has been observed that both strict lockdown and no lockdown measures can both contribute to postponement of attaining the peak of incidence rate evolution.

Parameter constant 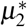 enables tuning the position of occurrence of the second wave component with respect to the first. The manifestation of the second wave component characterized by its center *µ*_2_ depends on the scale of community transmission that happend after non-medical interventions were implemented. In reality, it would be impossible for any country to trace, monitor and test a large segment of its population and ensure full compliance and enforcement of its policies across the entire geography. These segments of the population who got infected during the community transmission phase of SARS n-COV and who were untested untill the peak of the first wave component was attained, will manifest as the second wave component once testing becomes more widespread.

The constansts 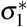 and 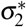 are control parameters that enables tuning the width of *σ*_1_ and *σ*_2_ respectively. A smaller the value of 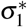 and 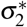 ranging between 0 and 1 indicate that Government strategies to manage the Covid-19 infection spread and the cooperation and compliance of its citizen resulted in successful flattening of the curve. Countries such as Australia, New Zealand, Taiwan, Hong Kong and South Korea have the smallest standard deviations of the first and second wave components and hence are considered to have implemented some of the successfull Covid-19 containment strategies. On the other hand, countries with larger values of *σ*_1_ and *σ*_2_ indicate that the pandemic containment strategies led to dealing with much higher incidence and mortality rates. The large standard deviations indicate the nature of variance in early implementation of control measures, testing rate for Covid-19, speed and effectiveness of contact tracing and importantly the compliance of the population adhering to health protocols and social distancing measures. The values of estimated parameters based on the selected constants described above are detailed in Table. 2.

**Table 2.** Details of the dual wave GLC parameters estimated based on Global Covid-19 incidence data as on May 31, 2020. NMI indicates Non-Medical Intervention (NMI) and NMI Delay Factor quantifies the delay in number of days to reach the peak amplitude of the first wave component. The GLC function parameters are estimated using equations 3 to 11 based on an iterative greedy selection of parameter constants.

|  | First Wave Component |  |  | Second Wave Component |  |  | NMI Delay Factor | Positivity Rate | Tests to Population Rate |
| --- | --- | --- | --- | --- | --- | --- | --- | --- | --- |
| | $h_1$ | $\mu_1$ | $\sigma_1$ | $h_2$ | $\mu_2$ | $\sigma_2$ | $t_\phi$ | | |
| Australia | 2679 | 68 | 9.71 | 297.68 | 85 | 24.29 | 0 | 0.57% | 4.89% |
| New Zealand | 525 | 74 | 8.46 | 87.5 | 86.3 | 16.91 | 0 | 0.58% | 5.42% |
| Taiwan | 144 | 64 | 13.71 | 32 | 89.6 | 27.42 | 0 | 0.62% | 0.30% |
| Hong Kong | 39.4 | 23.3 | 20 | 355 | 70 | 10 | 0 | 0.53% | 2.71% |
| South Korea | 4360 | 42 | 6 | 969 | 63 | 15 | 0 | 1.36% | 1.61% |
| Japan | 390.01 | 52.2 | 18.64 | 4291 | 87 | 18.64 | 0 | 6.10% | 0.21% |
| Norway | 1899 | 67 | 14.36 | 678.21 | 81.89 | 35.89 | 0 | 3.56% | 4.33% |
| Spain | 56038 | 71 | 14.79 | 32963.5 | 88.25 | 29.57 | 2 | 7.95% | 7.61% |
| Germany | 40856 | 73 | 15.21 | 18571 | 87.2 | 30.43 | 2 | 5.02% | 4.29% |
| France | 32616.1 | 72 | 13.11 | 58709 | 89 | 14.57 | 4 | 13.19% | 2.21% |
| Italy | 39554 | 68 | 13.71 | 26369 | 84 | 34.29 | 4 | 6.61% | 5.76% |
| Singapore | 7036 | 95 | 13.57 | 5412.31 | 112.27 | 33.93 | 0 | 10.86% | 5.04% |
| US | 221048 | 79 | 22.57 | 200953 | 93.36 | 90.29 | 0 | 11.29% | 4.54% |
| UK | 38896 | 87 | 17.79 | 35360 | 105.44 | 35.57 | 4 | 7.39% | 5.21% |
| Russia | 76873 | 113 | 23.79 | 64060.8 | 133.18 | 59.46 | 2 | 3.95% | 6.13% |
| Canada | 13454 | 95 | 23.4 | 8408.75 | 121 | 32.76 | 4 | 5.74% | 3.92% |
| UAE | 4084.67 | 100.64 | 28.11 | 6127 | 123 | 28.11 | 0 | 1.48% | 20.70% |
| India | 94039.5 | 160 | 38.57 | 62693 | 184.55 | 84.86 | 25 | 4.78% | 0.22% |
| Brazil | 252847 | 155 | 38.57 | 229861 | 179.55 | 84.66 | 20 | 50.04% | 0.35% |
| Sweden | 9694.5 | 163 | 42.43 | 4847.25 | 95.5 | 33.94 | 28 | 16.12% | 2.08% |

## Results and Discussion

The dual wave GLC function was used to model the incidence rate evolution trend for 20 countries based on the estimated parameters for both the first and second wave components. The results of the non-linear curve approximating the first and second wave components of weekly incidence rate trend are illustrated in Figures. 4 to 20. In order to compare the incidence rate trends in line with the non-medical intervention strategies implemented by each Government, the 20 countries are clustered into four groups. Cluster 1 represents countries that have successfully flattened the incidence rate curve. Similarily Cluster 2 represents countries that are on the verge of flattening the curve as on the current date of June 6, 2020. Cluster 3 represents countries that have reached the peaks and are on a decay trajectory. Finally, The Cluster 4 represents the set of countries that are yet reach their peak of incidence rate trend as on June 6, 2020. Based on the models generated for 20 different countries with diverse population dynamics, Government strategies it is established that the dual wave GLC function can approximate the incidence rate evolution of Covid-19.

**Figure 4.**
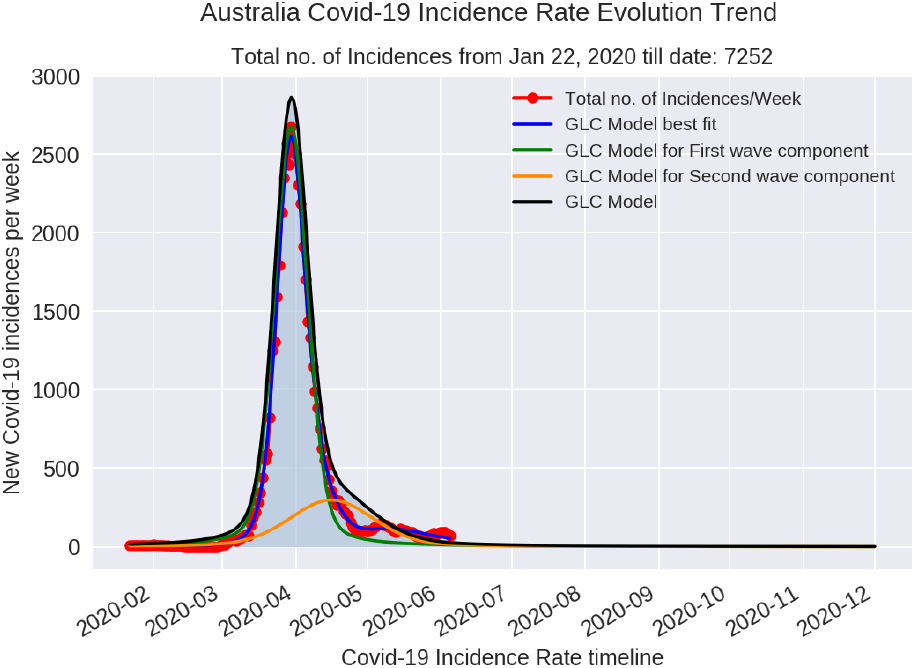
Dual wave GLC model for Australia

**Figure 5.**
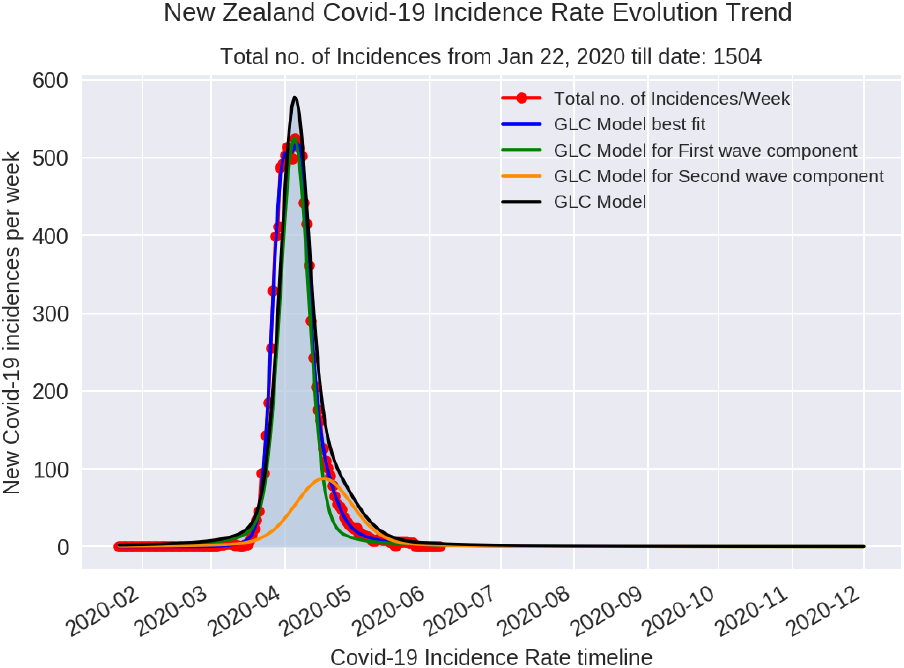
Dual wave GLC model for New Zealand

**Figure 6.**
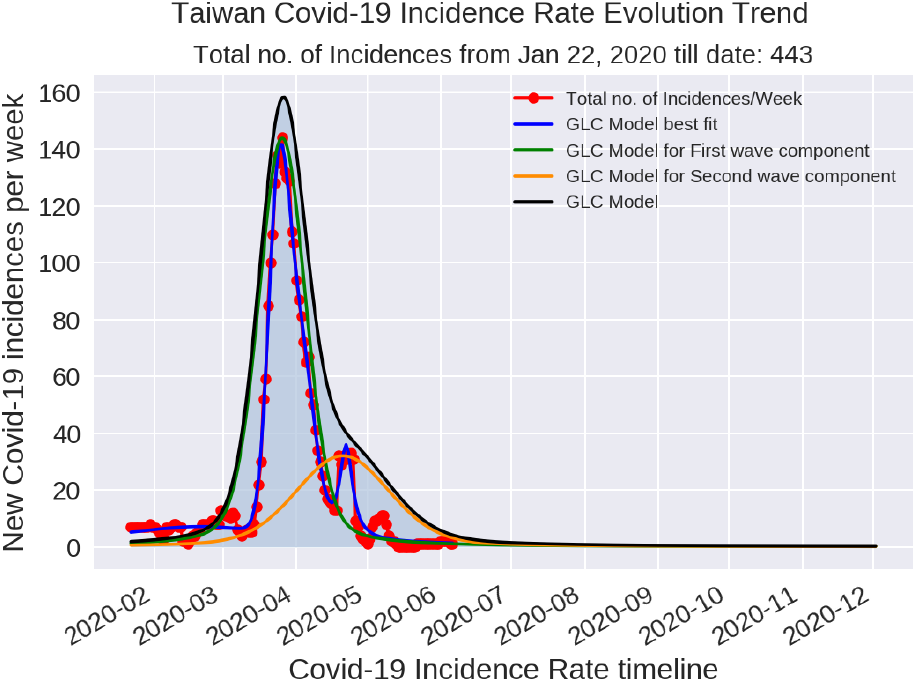
Dual wave GLC model for Taiwan

**Figure 7.**
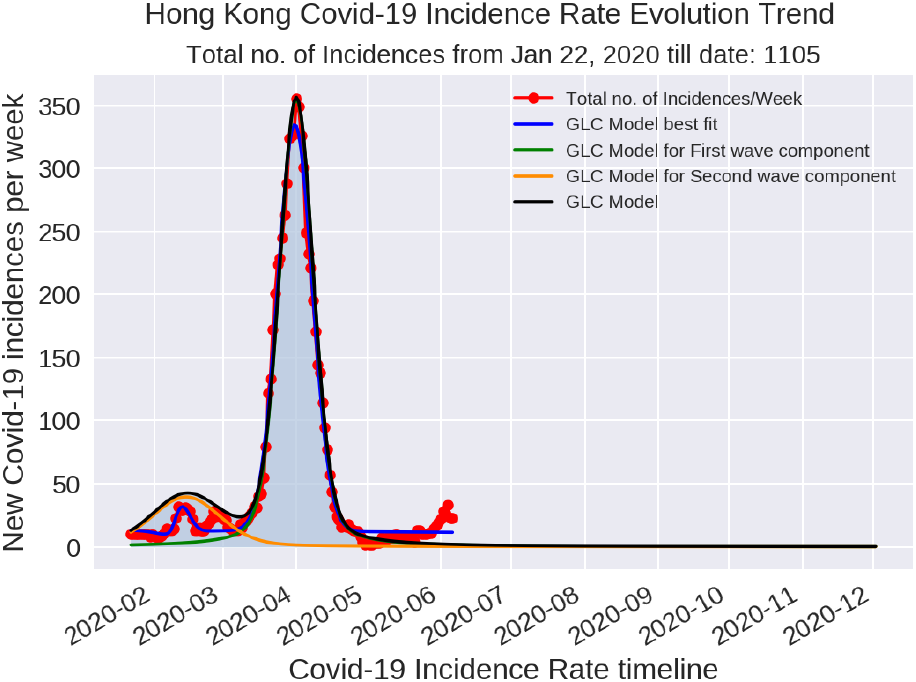
Dual wave GLC model for Hong Kong

**Figure 8.**
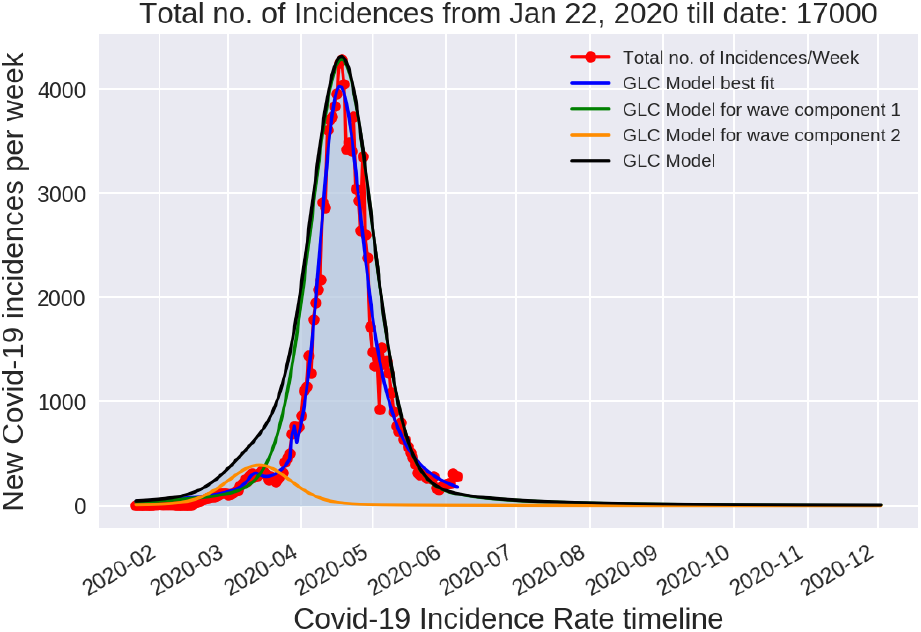
Dual wave GLC model for Japan

**Figure 9.**
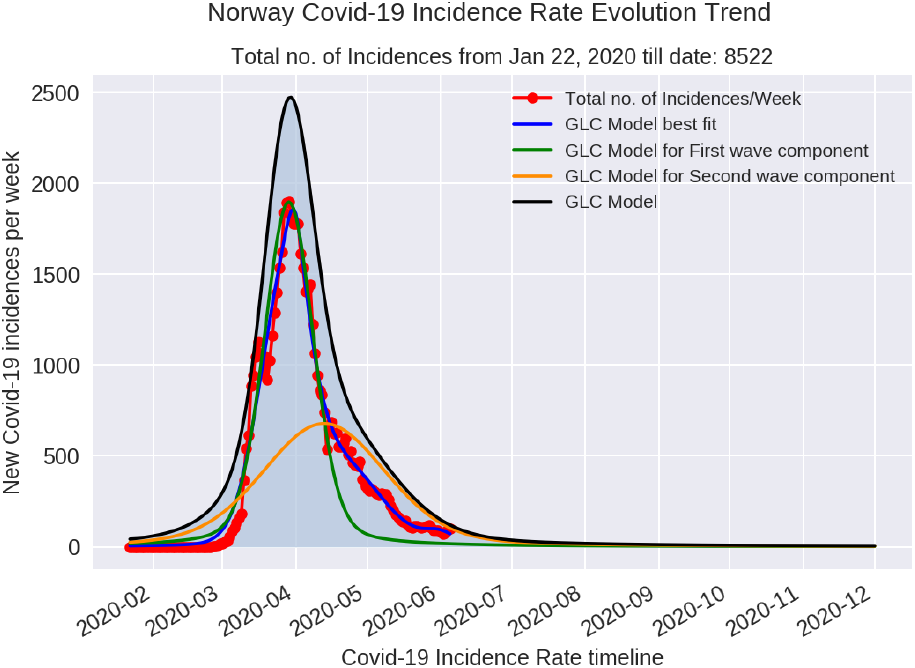
Dual wave GLC model for Norway

**Figure 10.**
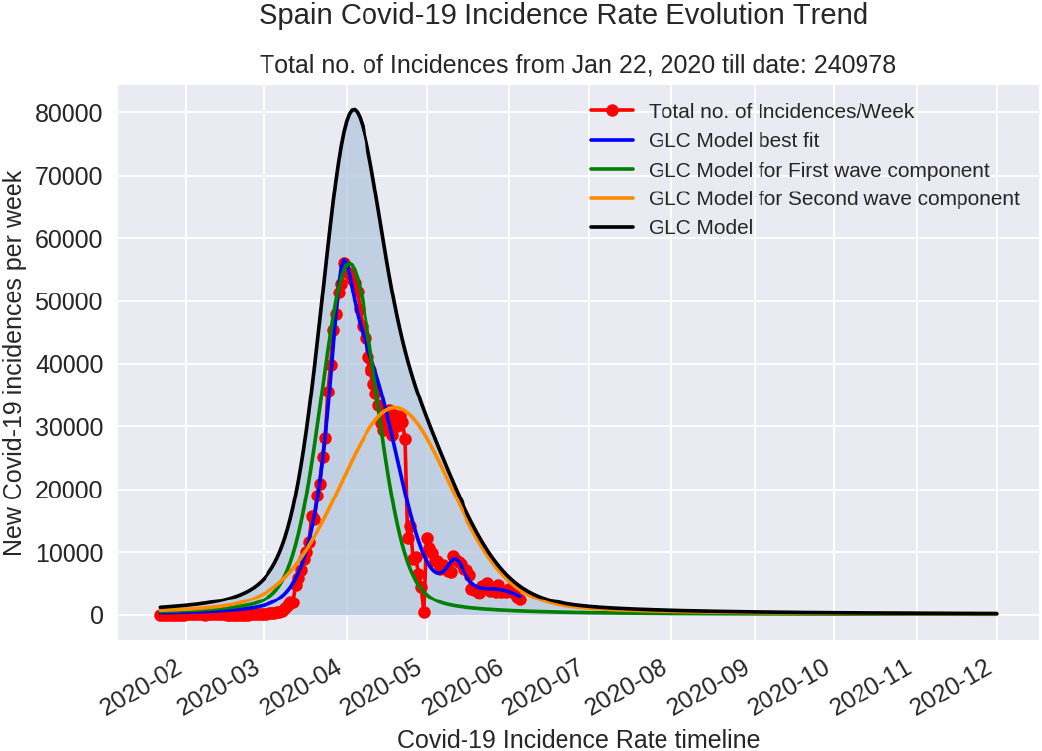
Dual wave GLC model for Spain

**Figure 11.**
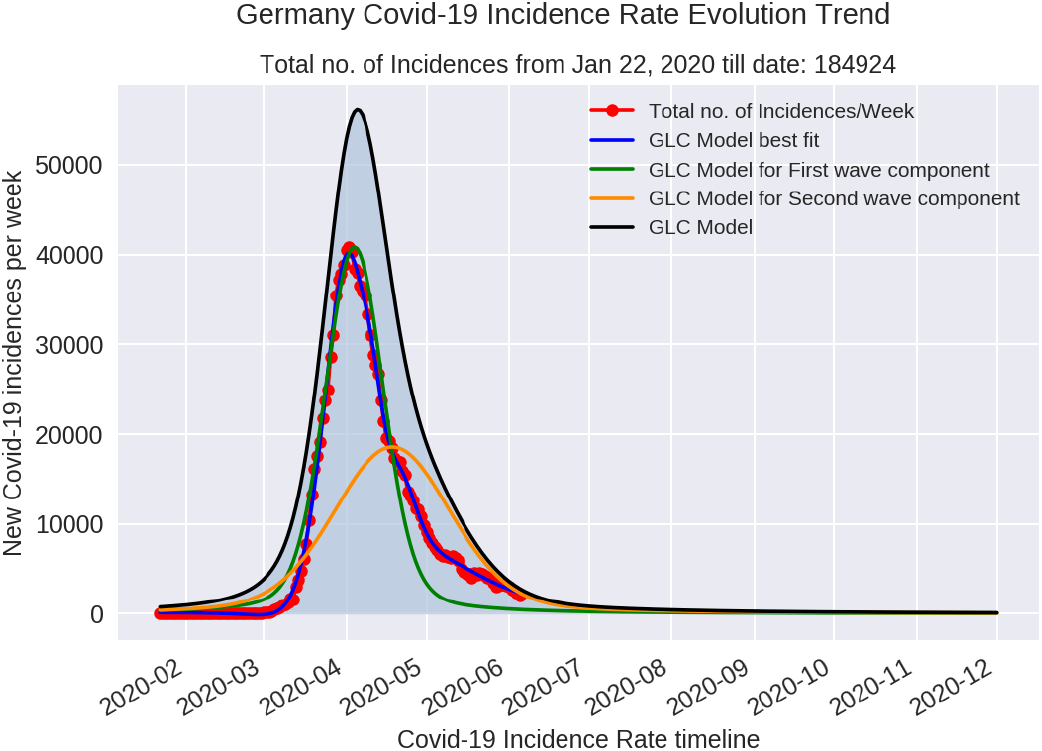
Dual wave GLC model for Germany

**Figure 12.**
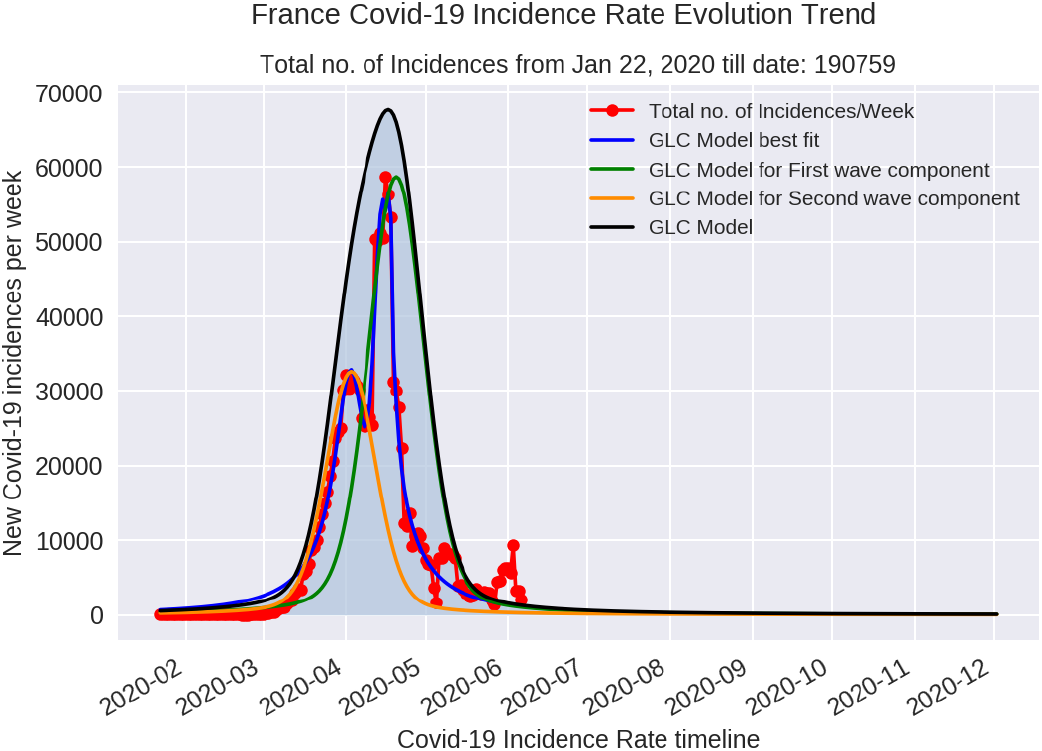
Dual wave GLC model for France

**Figure 13.**
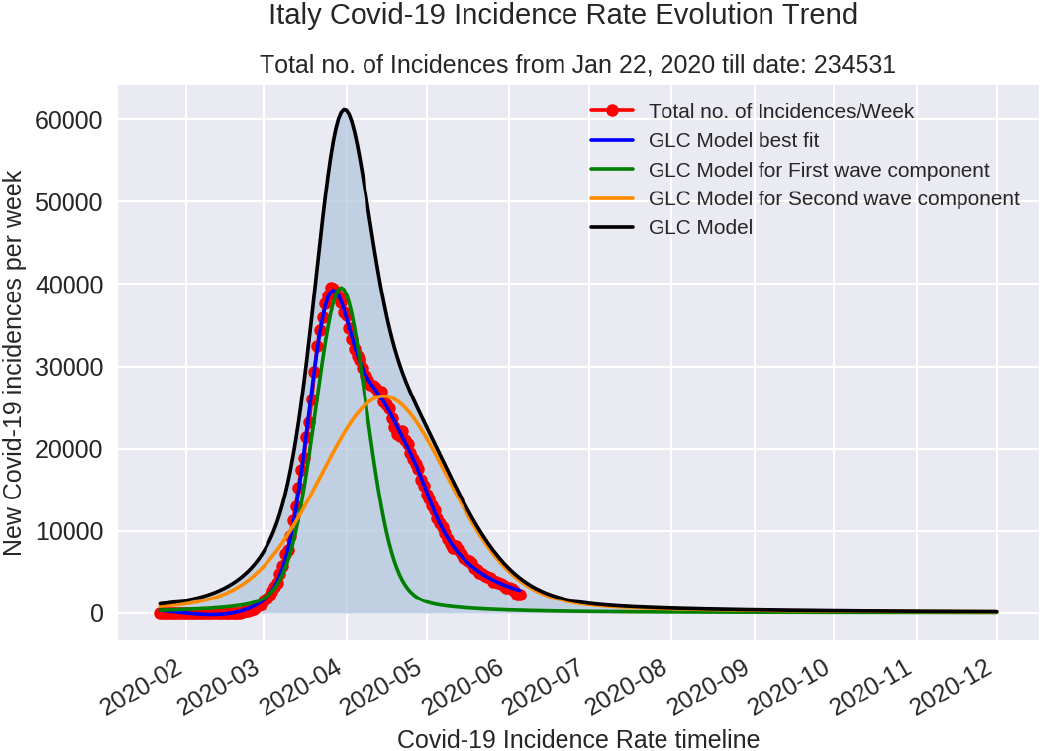
Dual wave GLC model for Italy

**Figure 14.**
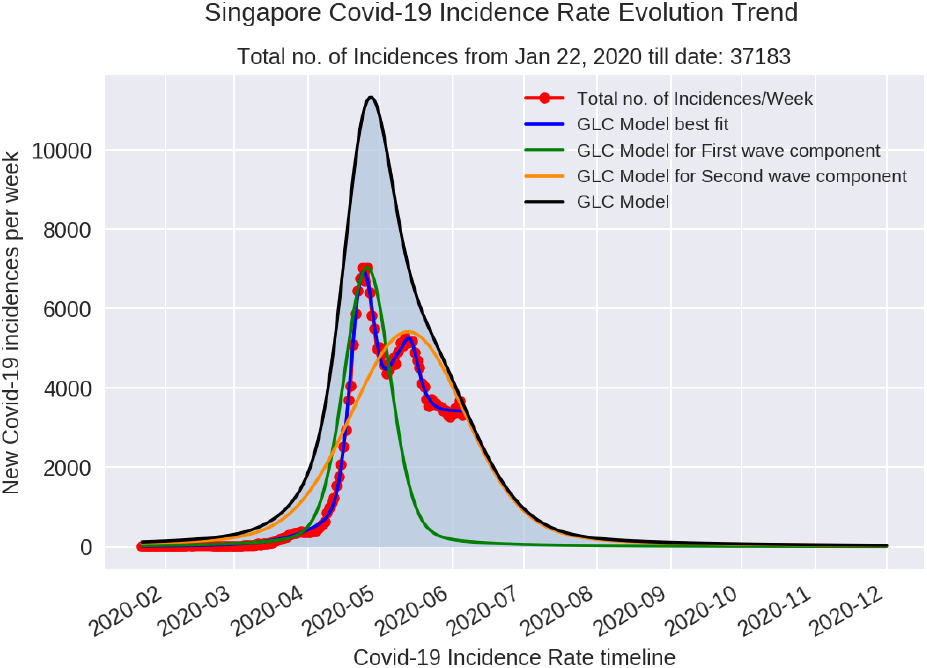
Dual wave GLC model for Singapore

**Figure 15.**
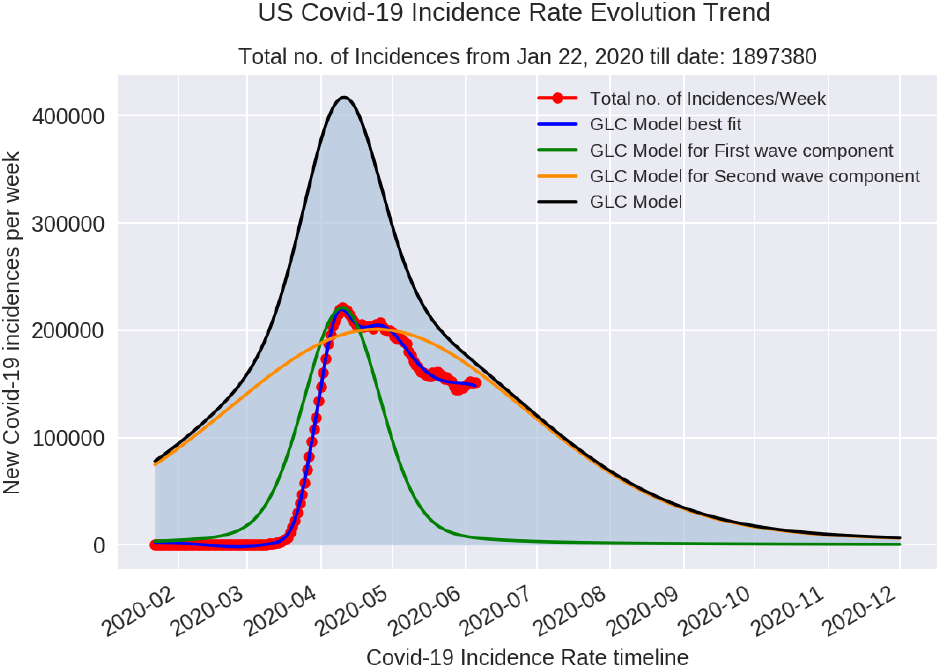
Dual wave GLC model for US

**Figure 16.**
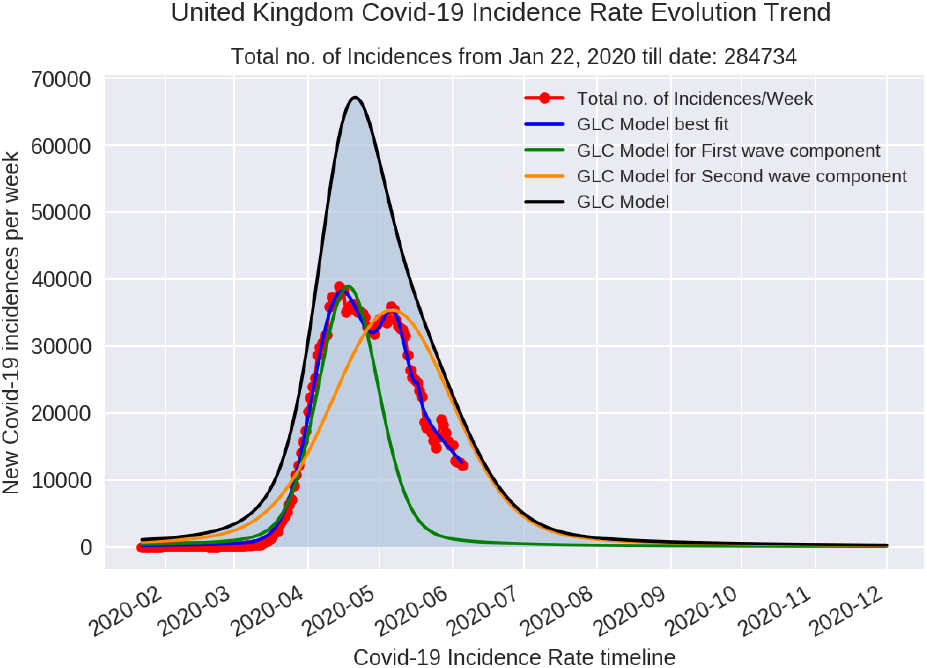
Dual wave GLC model for United Kingdom

**Figure 17.**
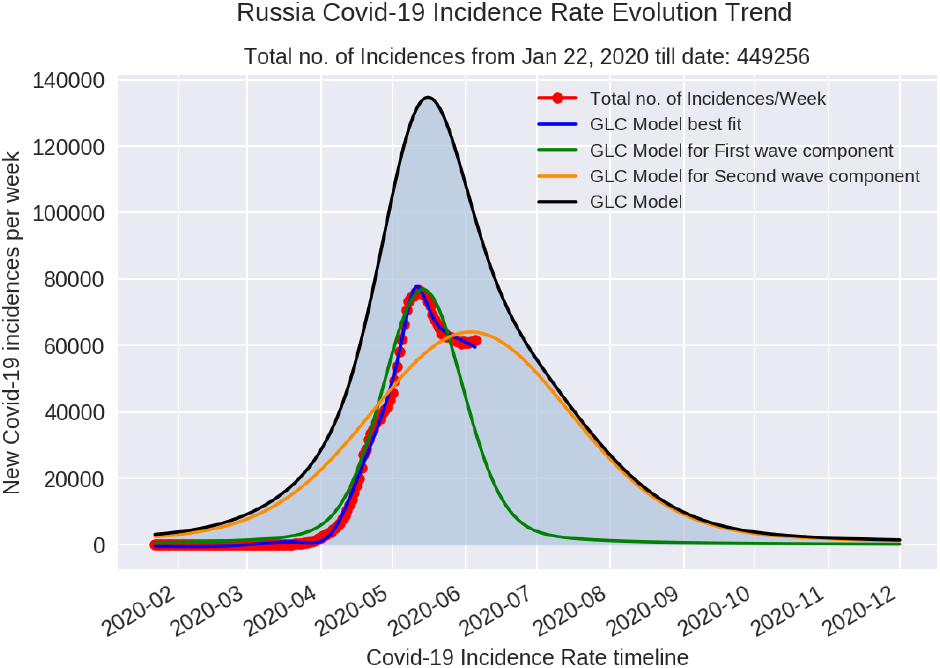
Dual wave GLC model for Russia

**Figure 18.**
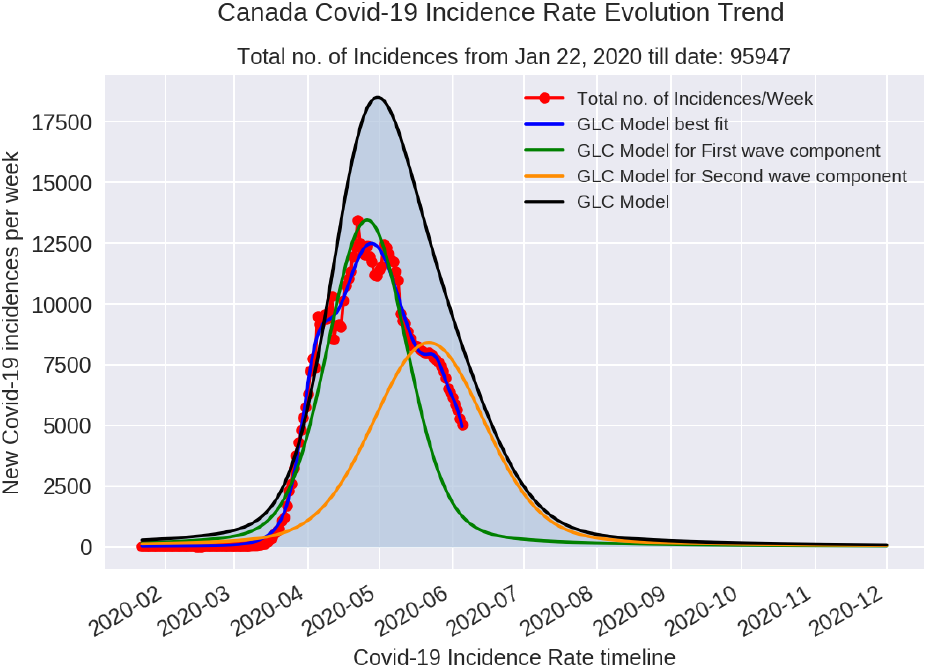
Dual wave GLC model for Canada

**Figure 19.**
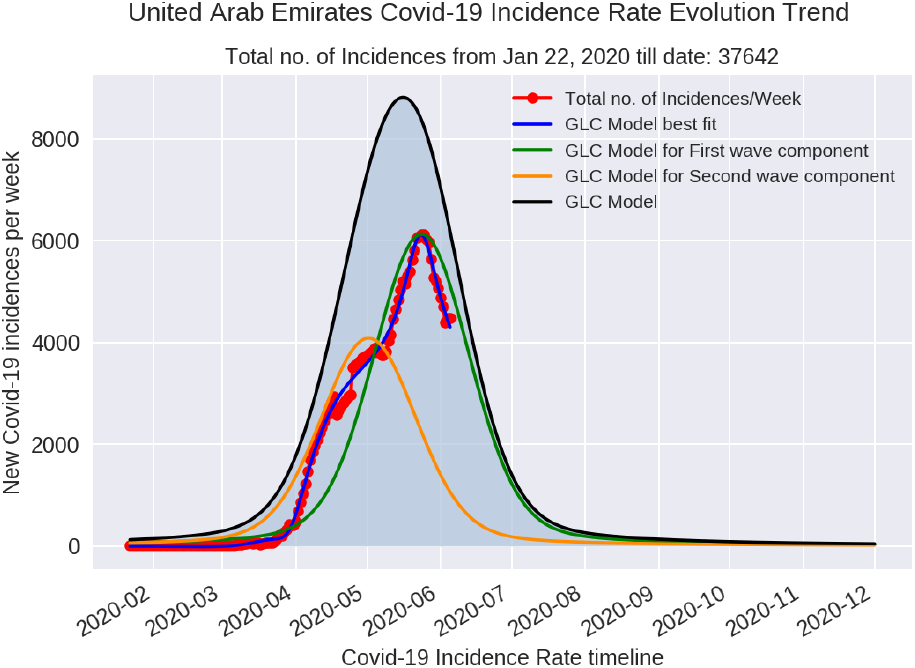
Dual wave GLC model for United Arab Emirates

**Figure 20.**
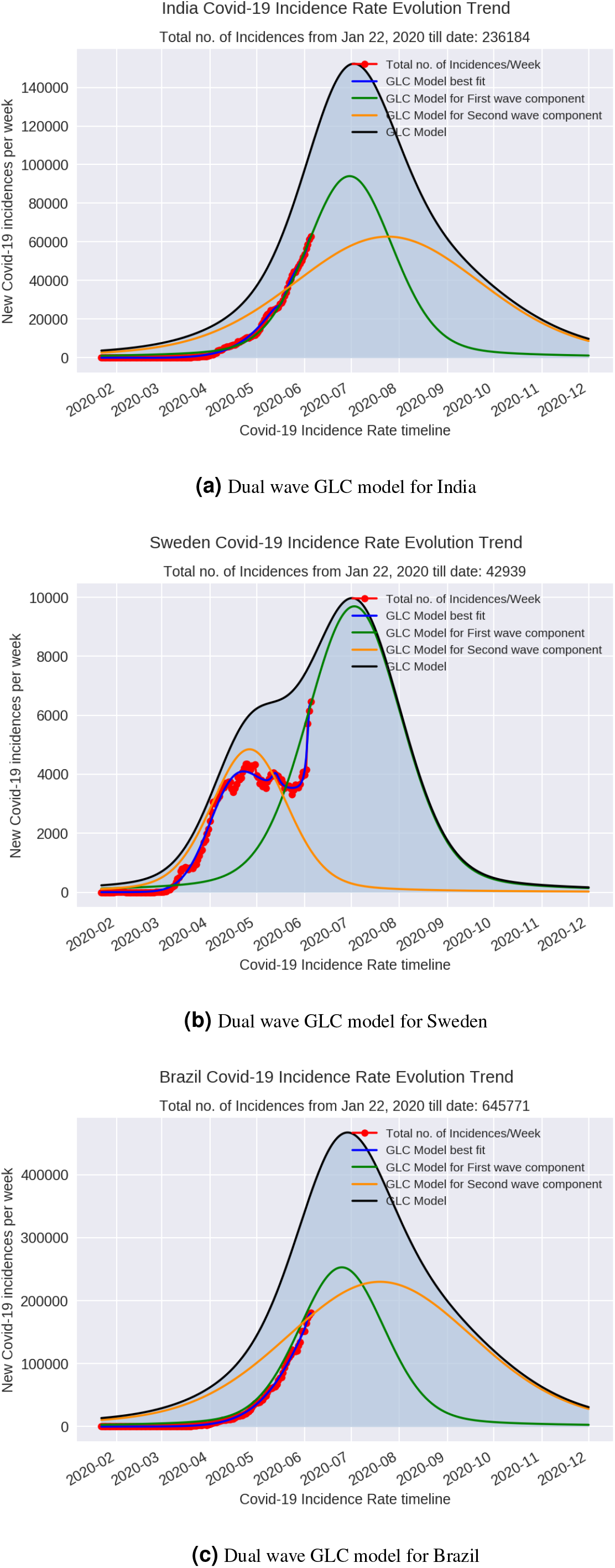
Dual wave GLC models for Cluster 4 countries

The probability density of the GLC curve can provide an estimate of the weekly incidence rate at that point in time. Given the nature of the Covid-19 pandemic in terms of the rate of its spread, incubation period, virulence, pathogen evolution and population response and compliance to Government health protocols, it would be impossible for any model to make a long term prediction of incidence rate trend. The same is applicable for the presented dual wave GLC model and hence a short forecast horizon of 180 days is considered for all the countries. Analysis of the various clusters described above shows a pattern of evolution of the incidence rates across geographies. The choice of parameter constants and correspondingly the estimated GLC model parameters on the success of different Covid-19 containment strategies by different Governments are detailed in Table. 1 and Table. 2

Referring to cluster 1, the GLC models for both first and second wave components indicate a smaller standard deviation as described earlier. This phenomenon indirectly gives insight into the coherence between Government-Citizen dynamics in successfully mitigating the spread of the pandemic. For example, in South Korea and Hong Kong citizens wore face masks right from the time their Governments stepped in with advisory on Covid-19 health protocols. Hence, one of the primary success metrics to “flattening of the curve” can be attributed to Government-Citizen coherence dynamics. Cluster 1 exhibits an example of positive coherence dynamics between the Government and its citizens. The other key metric to interpret the evolution of the incidence rate curve is the Covid-19 testing rate. Incidence rate data is fundamentally based on the scale of testing and positivity ratio. While it is important to test a population at the largest scale possible given the resources and capacity of a country, the fact that a tested individual can still be infected in the futrue still remains. Hence the positivity rate based on which the incidence rate numbers are reported is just a snapshot in time of the spread of SARS n-COV. This snapshot can be used to plan for the near future rather than to make absolute conclusions about a country’s containment strategy nor about the pathogen itself. Referring to Table. 2, it can be noted that the testing rates of Japan and Taiwan are the lowest in the world with testing done only on 0.21% and 0.30% of the populations respectively. However, the GLC model curves illustrate both Japan and Taiwan are among the few countries that succesfully flattened the curve with lowest incidence and mortality rates.

Since India, Sweden and Brazil have not yet reached their peak incidence rate, the GLC models for the three countries have been generated based on the average values of constant parameters 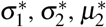 and 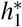 of first and second wave components from countries in clusters 1, 2 and 3, excluding outliers. The value of parameter constant 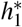 for the forementioned countries was selected based on the similarity of their incidence rate evolution curve compared to others in the various clusters. As of June 6, 2020, India’s incidence rate closely follows the trend of Russia whose weekly incidence rate peaked at 76873. Further, the only other country in the entire set of clusters which has a higher incidence rate trend compared to Russia is the US. However, India’s non-medical interventions is very different than that of US but comparatively similar to that of Russia. Accordingly, the 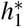 parameter constant for the first wave component for India is chosen to be 1.5. It means that the weekly incidences at peak for first wave component could reach approximately 91000 between June 15, 2020 and July 15, 2020. Considering this trend, India will potentially have approximately 50000 to 60000 weekly incidences at the beginning of September 2020.

The incidence rate trend for Sweden is still in the growth trajectory like India and Brazil. However, the main difference is that Sweden is currently having its second wave component being manifested. The growth trajectory of the second wave component as on June 6, 2020 is moving towards a higher amplitude compared to the amplitude of the first wave component. With a similar parameter constant selection strategy described above for India, Sweden could reach its peak weekly incidence rate between the July 1, 2020 and July 15, 2020. At this trend, the weekly incidence rate at the beginning of september could be approximately between 1500 to 2000. Brazil’s incidence rate trend has similarity that of US. Even with a very low testing rate of 0.35%, the positivity rate for Covid-19 is the highest in the world with 50.04%. This means 1 out of every 2 individuals tested confirms positive for Covid-19 as of testing statistics till May 31, 2020. With parameter constants similar to that of US, Brazil could potentially reach the peak weekly incidences of approximately between 240000 to 250000 if the country scales the testing efforts widely across the population. At the beginning of September 2020, the weekly incidence rate could still be around 170000.

The dual wave GLC function can be extended to a multiwave model to account for resurrgence of the Covid-19 incidences in the future. For example, South Korea has started witnessing the resurrgence of incidences after economic activity restarted from the first week of May 2020. Similarly, France and Hong Kong has seen resurrgence of incidences since the last week of May 2020. While extension of the dual wave GLC function is straigthforward to model resurrgence phenomenon, the multi-wave modeling is currently out of scope of this research.

Sensitivity analysis was performed on the GLC models generated for each country using Sobol Sensitivity analysis and Saltelli’s sample generating function [22, 23, 24]. Sobol’s sensitivity analysis decodes a model’s variance into composite variances related to the model parameters in progressive dimensions. A key outcome of the Sobol sensitivity analysis are the estimation of sensitivity indices that explain the variability of a model’s output based on individual parameter significance and the interactions between them. The Sobol Sensitivity Indices (SSI) represents first-order and total-order sensitivity which reflects the influence of an individual model parameters and the interaction between them [25]. Model parameters with *SSI >* 0.05 are considered significant and 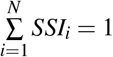. Further, the SSI of total-order indices is greater than SSI of first-order indices.

Figures 21, 22, 23 and 24 illustrate sensitivity analysis performed on Japan, South Korea, Germany and United Kingdom by generating 560 models each for a range of dual wave GLC model parameters. S1 and ST are the first-order and total-order sensitivity indices. S1 conf and ST conf are the bootstrap confidence intervals (1.96*standard error) and 95% confidence intervals. Higher the sensitivity indices, more critical are the model parameters for its output. For Japan, the parameter amp g 1 representing the Gaussian part of *h*_1_ in the first wave component is the most important parameter contributing to 85% of the model output variability followed by cen g 1 representing *µ*_1_ of the Gaussian part of the first wave component. SSI for South Korea indicates amp g 1 and cen l 1 contributing to approximately 37% each of the model output variability followed by cen g 1 and wid l 1 in the order of significance. For both Germany and UK, the parameter cen l 1 representing *µ*_1_ of the Lorentzian part of the first wave component contributes to more than 90% of the model output variability.

**Figure 21.**
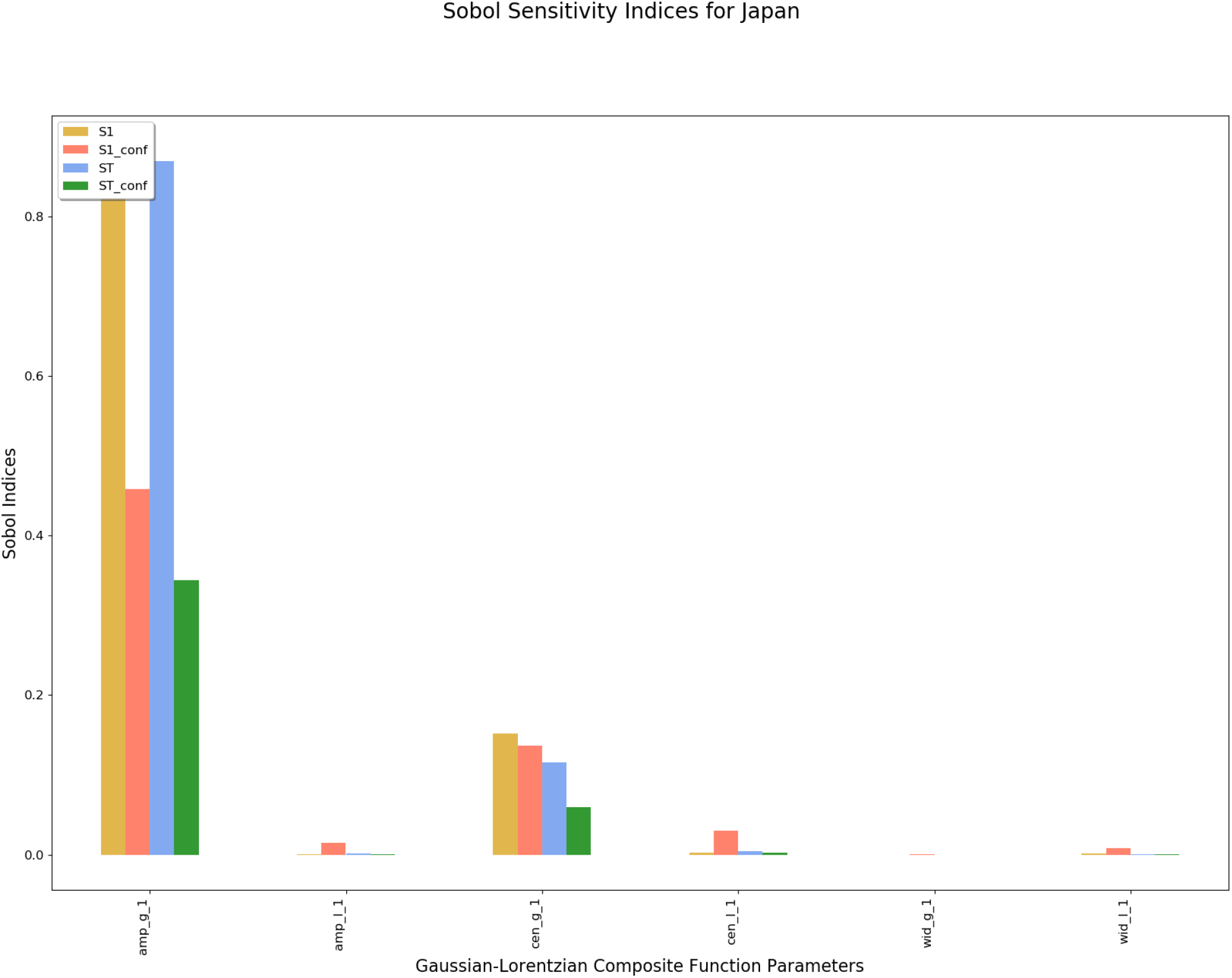
SSI for Japan

**Figure 22.**
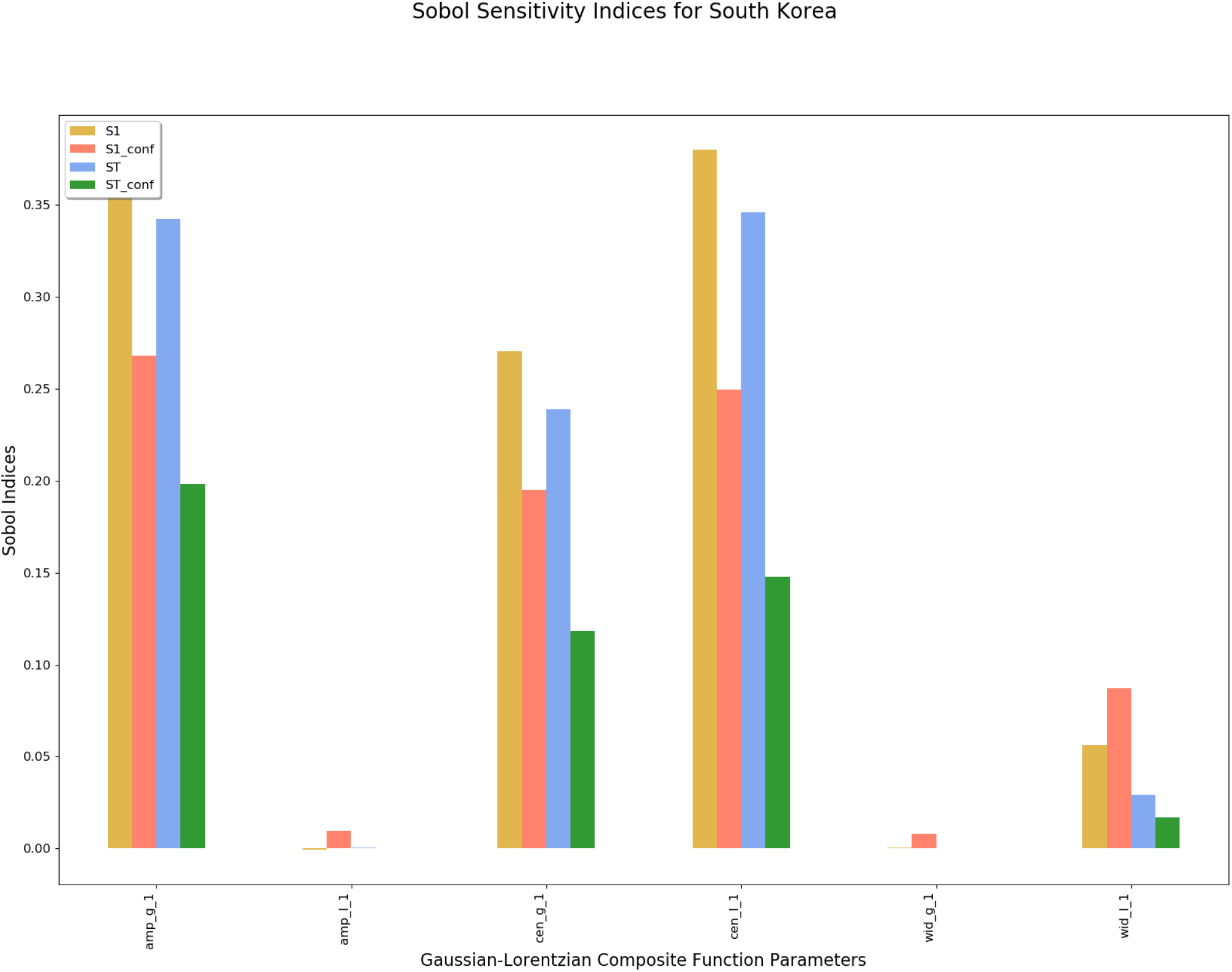
SSI for South Korea

**Figure 23.**
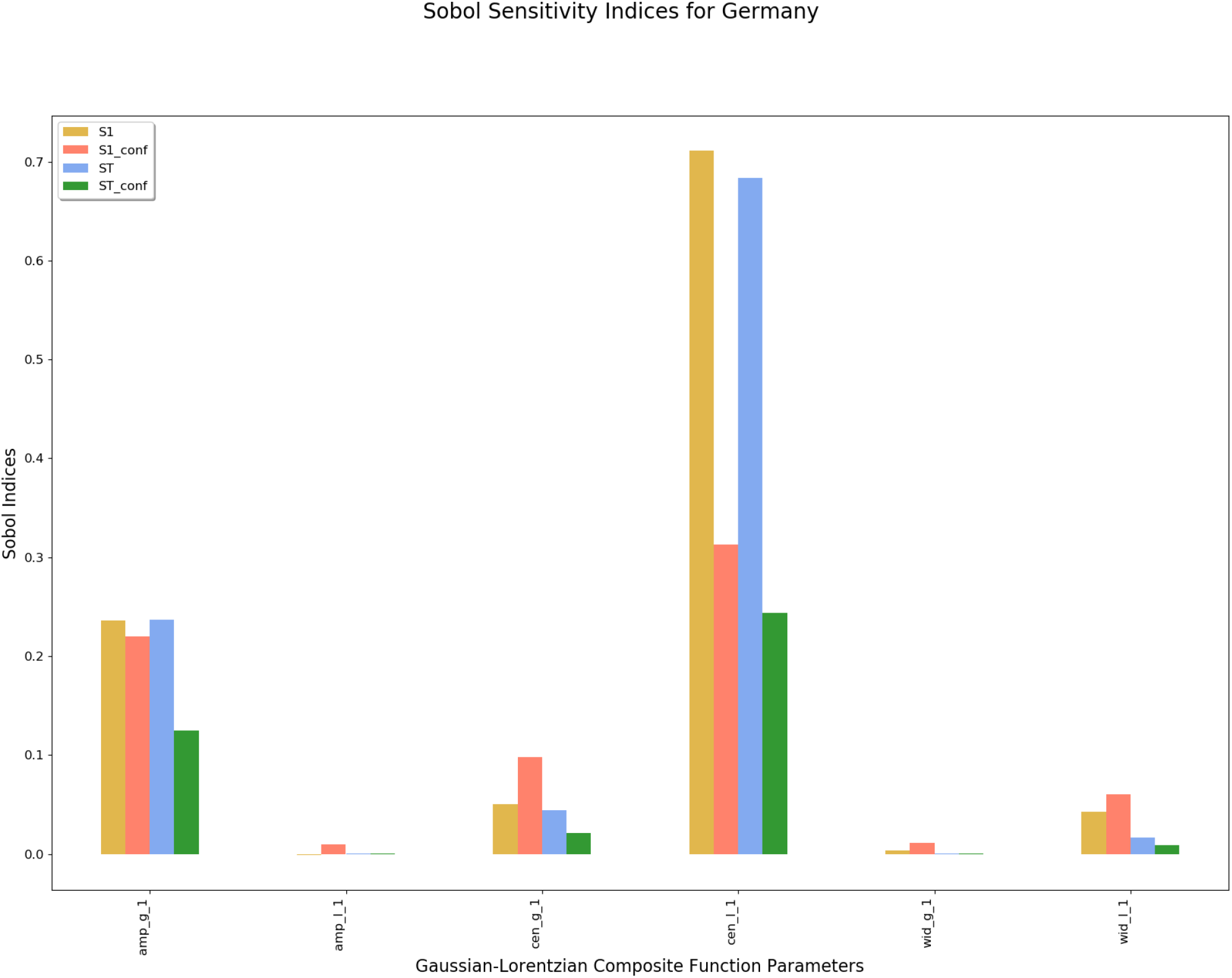
SSI for Germany

**Figure 24.**
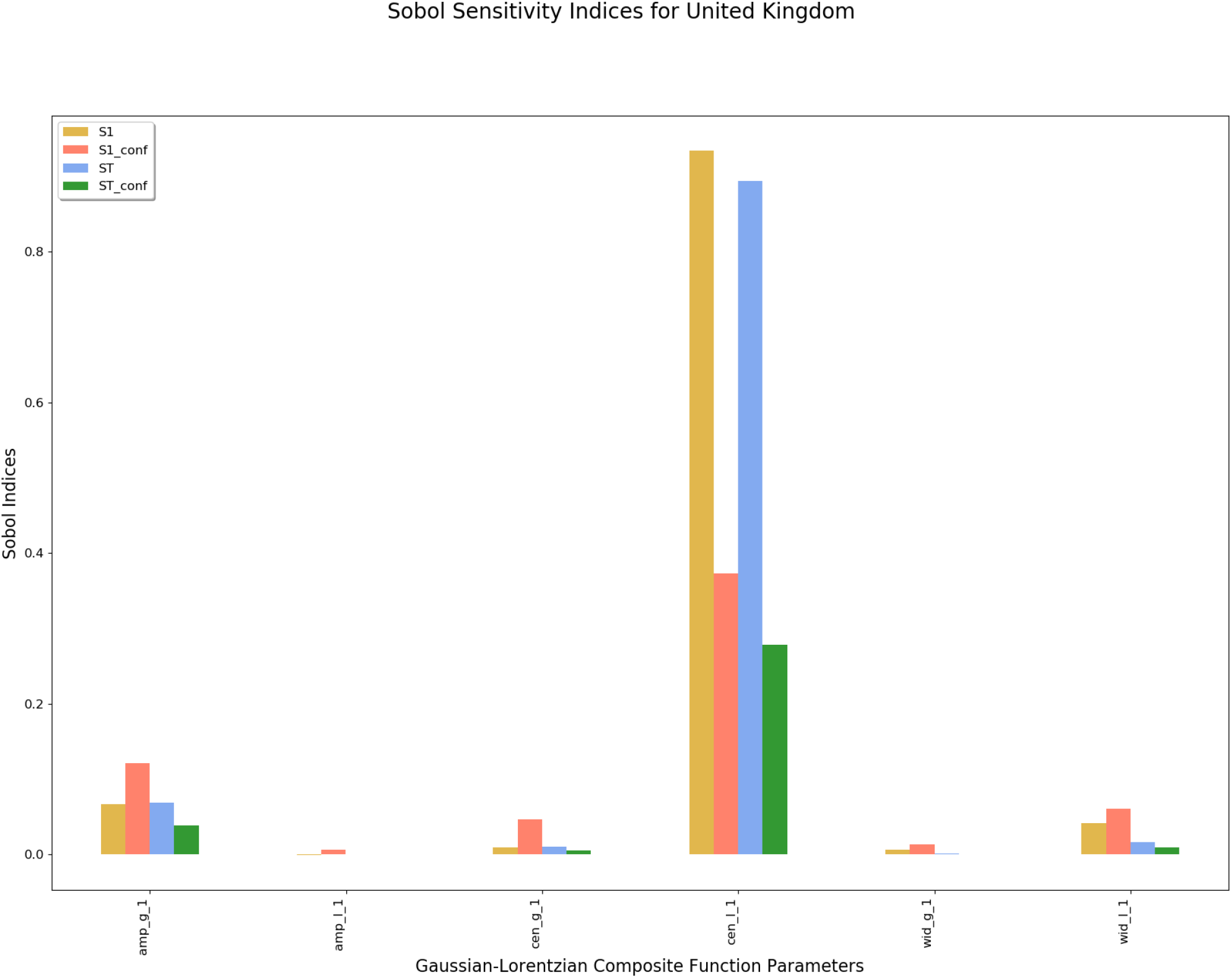
SSI for United Kingdom

In conclusion, the properties of Dual wave GLC model using Gaussian and Lorentzian functions has been shown to model the highly non-linear evolution trend of Covid-19 incidence rates. The mathematical properties of the GLC function can effectively be used to model the non-linearity of incidence rates and also account for the uncertainty in officially reported numbers. The resulting short term forecasts for each country especially India, Sweden and Brazil should be used only as a guidance in planned rebooting of the economies instead of absolute reliance on the predictions. Any mathematical model is as good as the data and the assumptions that come it. Modeling uncertainty is highly complex and hence any mathematical forecast need to be assessed along with experiential and ever evolving intuitive knowledge of pandemic.

## Data Availability

The data used for this research was obtained from the COVID-19 Data Repository by the Center for Systems Science and Engineering (CSSE) at Johns Hopkins University. Specifically, the time series dataset for global confirmed cases was used for the mathematical modeling.

https://github.com/CSSEGISandData/COVID-19/tree/master/csse_covid_19_data/csse_covid_19_time_series

## Data and Code Availability

Data is available publically from the COVID-19 Data Repository by the Center for Systems Science and Engineering (CSSE) at Johns Hopkins University. Code for dual wave GLC modeling can be obtained from https://bitbucket.org/radkris_xformics/covid19_glc.

## About the Author

Radhakrishnan Poomari is the Chief Data Scientist of Xformics Inc which is a startup head-quartered in Boston, Massachusetts, US. His primary focus is to identify unmeet needs in diverse domains and cross-pollinate domain knowledge with the state of the art AI, Machine Learning and first principles techniques to solve challenging problems.

Dual wave GLC Models for Cluster 1: Countries that flattened the curve

Dual wave GLC Models for Cluster 2: Countries that almost flattened the curve

Dual wave GLC Models for Cluster 3: Countries that are on decay trajectory from peak

